# A Pilot Study of a Ketogenic Diet in Bipolar Disorder: Clinical, Metabolic and Magnetic Resonance Spectroscopy Findings

**DOI:** 10.1101/2023.10.23.23297391

**Authors:** Iain H Campbell, Nicole Needham, Helen Grossi, Ivana Kamenska, Shane Sheehan, Gerard Thompson, Michael J Thrippleton, Melissa C Gibbs, Joana Leitao, Tessa Moses, Karl Burgess, Ben Meadowcroft, Benjamin P Rigby, Sharon A Simpson, Emma McIntosh, Rachel Brown, Maja Mitchell-Grigorjeva, Frances Creasy, John Norrie, Ailsa McLellan, Cheryl Fisher, Tomasz Zieliński, Giulia Gaggioni, Saturnino Luz, Harry Campbell, Daniel J Smith

## Abstract

**Background:** Preliminary evidence suggests that a ketogenic diet may be effective for bipolar disorder.

**Aims:** To assess the impact of a ketogenic diet in bipolar disorder on clinical, metabolic and brain magnetic resonance spectroscopy (MRS) outcomes.

**Method:** Euthymic individuals with bipolar disorder (N=27) were recruited to a 6-8 week single-arm open pilot study of a modified ketogenic diet. Clinical, metabolic and MRS measures were assessed before and after the intervention.

**Results:** Of 27 recruited participants, 26 began and 20 completed the ketogenic diet for 6-8 weeks. For participants completing the intervention, mean body weight fell by 4.2kg (p<0.001), mean BMI fell by 1.5kg/m^2^ (p<0.001) and mean systolic blood pressure fell by 7.4 mmHg (p<0.041). All participants had baseline and follow up assessments consistent with them being in the euthymic range with no statistically significant changes in symptoms (assessed by the Affective Lability Scale-18, Beck’s Depression Inventory and Young Mania Rating Scale). In some participants (those providing reliable daily ecological momentary assessment data; n=14) there was a positive correlation between daily ketone levels and self-rated mood (r=0.21, p<0.001) and energy (r=0.19 p<0.001), and an inverse correlation between ketone levels and both impulsivity (r =-.30, p<0.001) and anxiety (r=-0.19, p<0.001). From the MRS measurements, brain Glx (glutamate plus glutamine concentration) decreased by 11.6% in the anterior cingulate cortex ACC (p=0.025) and fell by 13.6% in the posterior cingulate cortex (PCC) (p=<0.001).

**Conclusions:** These preliminary findings suggest that a ketogenic diet may be clinically useful in bipolar disorder, for both mental health and metabolic outcomes. Replication and randomised controlled trials are now warranted.

**Study Registration Number:** ISRCTN61613198

## INTRODUCTION

### Ketogenic Diet: A Metabolic Treatment for Epilepsy

A ketogenic diet is established as a metabolic therapy for refractory epilepsy, supported by data from 13 randomized controlled trials and over a century of clinical use.(1,2) Preliminary case report, observational and pilot data have also suggested that a ketogenic diet may have beneficial effects in bipolar disorder.(3–6) Epilepsy and bipolar disorder may share some common pathophysiological mechanisms and several antiseizure medications are useful for bipolar disorder. Research into the effects of a ketogenic diet has highlighted mechanisms that may be relevant to bipolar disorder, including neuroprotective, anti-inflammatory, and positive metabolic effects.(7) We have previously proposed a mechanism of action of a ketogenic diet that may address deficits in brain cellular energetics (impaired insulin signalling and dysregulated glucose metabolism).(8,9)

### Cardiometabolic Health and Bipolar Disorder

Bipolar disorder carries a substantial metabolic burden, with high rates of obesity, type 2 diabetes, cardiovascular disease and reduced life expectancy.(10,11) Several current first-line medications for bipolar disorder increase cardiometabolic risk: adjunctive treatment strategies to ameliorate these effects are urgently needed.(12) A ketogenic diet has been demonstrated to improve cardiometabolic health outcomes within general population samples.(13) This pilot study was conducted primarily to assess the feasibility and acceptability of a ketogenic diet intervention in euthymic patients with bipolar disorder. These primary outcomes have been reported previously.(14) To inform the design of a future randomized controlled trial, we also assessed a range of secondary clinical, metabolic and brain magnetic resonance spectroscopy (MRS) outcomes, which are the focus of this report.

## METHODS

This was a single-group, non-randomised open interventional pilot study, with no control group. The study was registered at isrctn.com, with the registration number ISRCTN61613198, on 2nd March 2022. The authors assert that all procedures contributing to this work comply with the ethical standards of the relevant national and institutional committees on human experimentation and with the Helsinki Declaration of 1975, as revised in 2013. All procedures involving human subjects/patients were approved by the South East Scotland Research Ethics Committee 02 (REC ref 22/SS/0007) and NHS Lothian Research and Development. The Academic and Clinical Central Office for Research and Development (ACCORD) provided sponsorship. Written informed consent was obtained from all participants.

### Study Participants

Recruitment for study participants began on 27th April 2022 and was carried out in collaboration with the charity Bipolar Scotland, via local support groups, social media and the Bipolar Scotland newsletter.

#### Participant Eligibility Criteria

Participants were individuals diagnosed with bipolar disorder, according to DSM-IV criteria. Eligibility was contingent upon a period of clinical euthymia lasting at least 3 months, defined by an absence of clinically significant depressive or hypomanic/manic episodes. The age range for participation was 18–70 years. Understanding of English language and residence in Scotland were also required.

#### Initial Exclusion Criteria

The initial exclusion criteria excluded participants with conditions including pregnancy, breastfeeding, intention to conceive within 3 months, active substance misuse, recent adherence to a ketogenic diet (within the past 2 months), adherence to a vegan diet (vegan ketogenic diet is widely used but our team did not have experience delivering this), recent hospital admission (within the past 3 months), concurrent participation in other research studies, inability to complete baseline assessments, and a history of liver, kidney, or cardiovascular diseases. Severe hyperlipidaemia, indicated by familial hypercholesterolaemia or a total cholesterol level exceeding 7.5 mmol/L, was also an exclusion criterion.

#### Additional Exclusion Criteria Applied During the Study

As the pilot study progressed, further exclusion criteria were added to address emergent considerations. These were a diagnosis of diabetes, engagement in activities requiring a high energy expenditure (such as long-distance running), and significant recent changes to psychotropic medication use.

### Participant Assessments and Data Collection

#### Screening and Baseline Procedures

Participants were briefed about the study’s objectives and the eligibility criteria were assessed. On successful screening and consent, baseline assessments were conducted and participants were provided with comprehensive guidelines on adopting and adhering to a ketogenic diet. This included information on potential risks and detailed instructions for diet monitoring. Participants were also required to submit a 3-day food diary and a pre-ketogenic diet information sheet in advance to facilitate the customisation and planning of their diet by the study dietitian.

#### Assessment and Monitoring

Assessments were conducted at baseline and during the follow-up period (6–8 weeks). Baseline assessment included a review of medical and medication histories, measurements of blood pressure and body mass index (BMI), and completion of a diagnostic interview. Participants also completed several mental health symptom scales, including the Affective Lability Scale 18 (ALS-18), Beck’s Depression Inventory (BDI), and the Young Mania Rating Scale (YMRS). Quality of life was assessed using the Within Trial Resource Use Questionnaire, the EuroQol 5D, and the Work Productivity and Activity Impairment Questionnaire, tailored to capture data on health and social care resource utilisation, household expenditure on food and beverages, and information on employment and absenteeism. Fasting venepuncture and MRS brain scans were conducted pre- and post-intervention. Post-analysis of the study data it was ascertained that one participant had not fasted on the morning of their baseline appointment for venepuncture and MRS. Their data did not present as an outlier for any variable or significantly alter the findings of the study and so remains in the analysis. Data collection was facilitated through both paper questionnaires and a secure online platform, with face-to-face interactions occurring at designated clinical and imaging facilities.

#### Brain Magnetic Resonance

Brain MR imaging and spectroscopy were acquired using a 3-T clinical MRI scanner (MAGNETOM Prisma, Siemens Healthcare, Erlangen, Germany) with a 32-channel receive head coil. A 3D T1-weighted image (T1w; TR/TE/TI = 2500/4.37/1100 ms, flip angle = 7°, 1.0-mm isotropic resolution) was acquired to facilitate placement of MRS voxels covering the anterior cingulate cortex (ACC; 20 x 20 x 20 mm3, 128 averages, acquisition time 4m26s), right dorsolateral prefrontal cortex (RDLPFC; 30 x 15 x 15 mm3, 96 averages, acquisition time 3m22s) and posterior cingulate cortex (PCC; 20 x 20 x 20 mm3, 128 averages, acquisition time 4m26s). The ACC and RDLPFC were selected as regions of interest (ROIs) based on previous literature implicating these regions in the pathophysiology of bipolar disorder,(15,16) whereas the PCC was selected to generate optimal data quality. At follow up, voxels were automatically placed at the same location using the AutoAlign feature. MRS was obtained using point-resolved spectroscopy (PRESS; TR/TE = 2000/30 ms, flip angle = 90°, receive bandwidth 4000 Hz, 4096 data points, with a single transient acquired without water suppression). Please see supplementary materials for further details.

#### Daily Monitoring and Rest/Activity Pattern Tracking

Participants engaged in daily self-monitoring of glucose and ketone levels using a KetoMojo device, initially reporting readings via text and later through bluetooth-enabled app syncing. Continuous actigraphy was deployed over a nine-week period to monitor rest/activity patterns, using 50hz AX3 actigraph devices worn in succession for three weeks each. GGIR software was used for accelerometer data processing. Daily ecological momentary assessments (EMA) captured changes in anxiety, mood, energy levels, impulsivity, and speed of thought, initially via text messages and subsequently via an online tool.

#### Post-intervention Evaluation

Following the intervention, semi-structured telephone interviews were conducted with 15 participants and four research clinicians involved in the study to facilitate a process evaluation, the findings of which are reported separately (17).

#### Dietary Intervention Protocol

The intervention was a modified ketogenic diet, with a macronutrient distribution of 60–75% calories from fats and 5–7% from carbohydrates, with the remainder sourced from protein. In consideration of blood cholesterol and triglycerides, a preference for unsaturated fats was advised. Adjustments to individual macronutrient ratios were made throughout the study by the study dietitian based on various factors, including the attainment of ketosis defined by a target blood ketone level of 1–4 mmol/L and glucose levels of 4–7.8 mmol/L. In cases where weight loss was both desired and considered safe, a caloric deficit was prescribed to promote body fat as an alternative ketone source. Participants engaged in the diet for a 6–8 week duration, which included an initial adaptation phase, alongside continued standard medical care from their regular healthcare providers.

#### Dietary Support and Management

Throughout the intervention, participants had weekly remote consultations with a dietitian, supplemented by additional contacts as needed. Participants received guidance to ensure understanding of the diet’s principles, support in adhering to the diet, assistance with troubleshooting dietary challenges and management of potential side effects. The diet plans specified total caloric and macronutrient (in grams) daily intake, portioned accordingly. Customized recipes were provided to align with individual dietary needs. Supervision was continued throughout the diet’s cessation phase, with support offered to those opting to maintain the dietary changes. Behavioural strategies were integrated to enhance diet adherence, including bi-weekly adherence checklists guided by the COM-B framework (which highlights the influence of capability, opportunity, and motivation on behaviour). These have been analysed in a separate process evaluation.(17)

#### Nutritional Supplementation and Monitoring

Consistent with international nutritional guidelines, participants were counselled on increasing fluid intake and supplementing their diet with a broad-spectrum multivitamin and mineral supplement, along with a calcium and vitamin D supplement. Participants were also educated on managing possible adverse effects, such as hypoglycemia or hyperketosis. Pre-intervention and follow-up health screenings, including analyses of urea and electrolytes, liver function tests, and lipid profiles, were mandatory for all participants. These were carried out to rule out significant hepatic or renal dysfunction or familial hypercholesterolemia and to monitor for adverse changes related to diet, such as alterations in liver function or lipid levels.

#### Primary and Secondary Outcomes

The primary outcomes of this study assessing feasibility, acceptability and safety have been reported in a previous publication. (14) Here we report the secondary outcome measures (ISRCTN61613198):

1. Mood stability measured by the Affective Lability Scale (ALS18) and depression by the Beck Depression Inventory (BDI) at baseline and at 8-week follow-up;
2. Hypomania/mania symptoms measured using the Young Mania Rating Scale (YMRS) at baseline and at 8-week follow up;
3. Glucose/ketone ratio measured using Ketomojo devices daily throughout the 10-week study period;
4. Identification of specific metabolic changes in glucose, ketones and tricarboxylic acid (TCA) metabolites associated with the ketogenic diet, measured using serum and brain MRI measures at baseline and at week 8;
5. Mood, energy, speed of thought, impulsivity and anxiety measured using visual analogue scales from 1-100 on daily EMA assessments during the 10 week study period;
6. Episodes of bipolar depression and hypomania/mania occurring during the study period assessed by the MINI Neuropsychiatric assessment at baseline and at week 8;
7. Sleep duration and circadian activity/rest rhythmicity parameters, assessed by actigraphs throughout the 10-week period (to include 2 weeks of stepping down the diet).

### Data Management and Statistical Analysis

#### Clinical and Metabolic Outcomes

The statistical analysis used standard methods in SPSS and Excel software (Windows PC version). We calculated mean and median values, standard deviations, and the change from baseline to follow-up for metabolic parameters, BDI, ALS-18 and YMRS values. We used the paired samples t-test function in SPSS to compare the mean at baseline and follow up and determine statistical significance (defined as p <0.05). Available case analysis of all participants who had baseline and follow up measures was performed and the sample size for each variable are given in the results. HOMA-IR was calculated from fasting insulin and glucose levels using the standard equation. (18)

#### Ecological Momentary Assessment and Analysis

Text messages were used for daily EMA assessment of mood, energy, speed of thought, impulsivity, and anxiety. During the study, the initial EMA rating instructions delivered to participants 1-12 were highlighted by participants as difficult to follow due to inconsistent individual interpretations of normal and pathological ranges on the 100 point scale (e.g., in the mood category participants chose differing ranges between 1-100 to indicate their healthy mood, depression and hypomania, making comparisons unworkable). Thresholds for normal ratings and pathological ratings of depression and (hypo)mania were therefore added to the instructions for patients 13-27 (see supplementary material). Note that EMA analyses were performed only on this second sub-group (n=14).

### Brain Magnetic Resonance (MR) Imaging and Spectroscopy

MRS data were processed using Osprey version 2.4.0,(19) including frequency alignment and phasing of transients, co-registration to the T1w anatomical image and segmentation of tissues within the voxels. Data were fitted in the frequency domain using a linear combination modelling approach, including default basis functions for lipid, macromolecular resonances and metabolites (ascorbate, aspartate, creatine (Cr), gamma-aminobutyric acid, glycerophosphorylcholine (GPC), glutathione, glutamine (Gln), glutamate (Glu), myo-inositol (mI), lactate, N-acetylaspartate (NAA), N-acetylaspartylglutamate (NAAG), phosphocholine (PCh), phosphocreatine (PCr), phosphoethanolamine, scyllo-inositol and taurine). Of the metabolites, tNAA (NAA + NAAG), Glx (Gln + Glu), tCr (PCr + Cr), tCho (PCh + GPC), mI, Glu and Gln were quantified with sufficient reliability for inclusion in further analysis and are reported here. Metabolite levels were quantified as water-scaled, relaxation-corrected and tissue-corrected estimates of molal concentration.(20) Spectra were automatically excluded if either the Cr or water resonance linewidth (full width at half maximum) was equal to or greater than 0.1 ppm; in addition, spectra and model fits were inspected visually and excluded in case of poor model fit, excessive lipid contamination, spurious signal contamination, low signal, poor water suppression or baseline distortion.

#### Metabolomics

Fasting blood samples were taken at baseline appointments and at 6-8 week follow-up appointments. Global metabolomics analysis was performed on a total of 36 serum samples, corresponding to 18 participants who gave a baseline and follow up blood sample and included one unfasted participant. Standard biochemical parameters included HbA1c, C-reactive Protein (CRP), beta-hydroxybutyrate (BHB), insulin, glucose and lipid levels. A more detailed exploratory investigation of untargeted metabolomics was also performed using Rapid HILIC-Z Ion mobility mass spectrometry (RHIMMS) analysis to probe the serum metabolome, annotating a total of 358 metabolic features. Please see supplementary materials for further details.

## RESULTS

The primary outcomes for this study (acceptability and feasibility) have been published previously: 20 out of 26 participants who started the diet adhered to the intervention and completed the outcome assessments.(14)

### Clinical Outcomes

All participants were clinically euthymic at baseline and median scores at week 8 remained within euthymic range, with no statistically significant changes: ALS 15 (IQR = 16.5) to 12.5 (IQR = 13.25) (p = 0.80) (n =18); YMRS 0 to 0 (p = 0.86) (n =20) BDI 8.5 (IQR = 11) to 9 (IQR = 9.25) (p = 0.55) (n=18). The MINI Neuropsychiatric Interview assessments reported one episode of hypomania during the study period.

Daily EMA measurements of mood, energy, speed of thought, impulsivity and anxiety on a subset of participants who provided reliable EMA data (n=14) were plotted against daily ketone levels (Figure 1).

**Figure 1.**
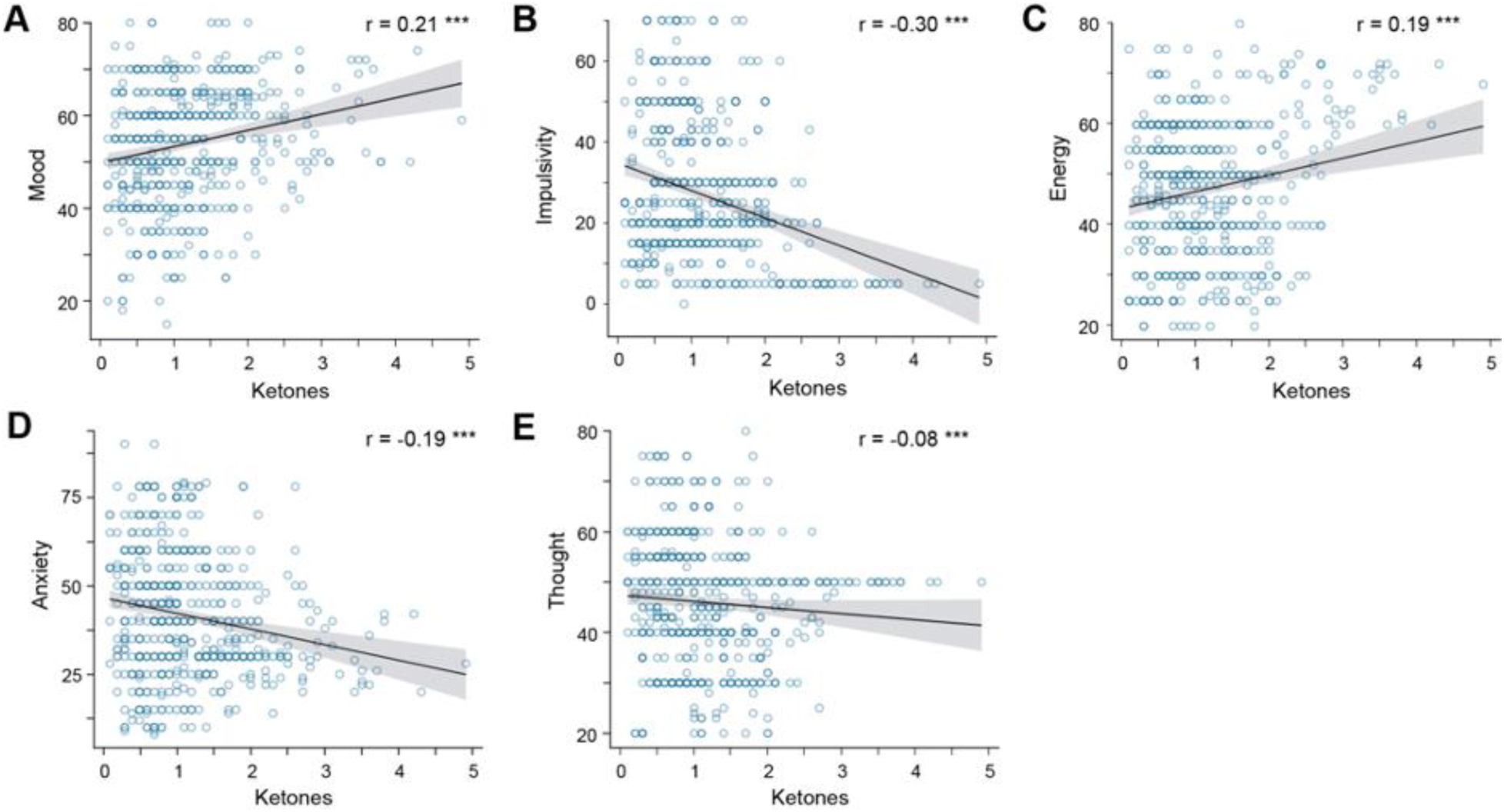
Daily Ecological Momentary Assessment and Ketone Levels (for a sub-set of n=14 participants)

A positive correlation was observed between daily ketone level and mood (r = 0.21, p < 0.001) and energy (r = 0.19 p < 0.001) scores, with a negative correlation between ketone levels and both impulsivity (r =-.30, p < 0.001) and anxiety (r =-0.19, p < 0.001). No correlation was observed with speed of thought (r = -0.08 p > 0.05) (Fig.1). 7 of the 20 participants who completed the intervention requested not to participate in the dietary cessation period and opted to continue on a ketogenic diet.

### Metabolic Outcomes

At baseline, 70.3% of participants were obese or overweight. Mean body weight decreased by 4.2kg (from 81.2 ± 13.2kg to 77.0 ± 11.7kg, p < 0.001) (n = 18). Mean BMI decreased by 1.5 kg/m^2^ from 28.2 ± 4.2 kg/m2 to 26.7 ± 3.9 kg/m2 (p < 0.001) (n = 20). Mean systolic blood pressure decreased by 7.4mmHg from 133.1 ± 16.2 mmHg to 125.7 ± 11.9 mmHg (p = 0.041) (n = 19).

The changes in mean values of the standard biochemical parameters (reported below as mean values at baseline then at follow up) were not statistically significant: mean diastolic blood pressure 81.5 ± 12.20 mmHg to 77.4 ± 7.7 mmHg (p = 0.162) (n = 19); fasting insulin 75.3 ± 44.0 to 68.4 ± 49.0 pmol/L (p = 0.540) (n = 17); fasting glucose 5.0 ± 0.6 to 4.7 ± 0.4 mmol/L (p = 0.12) (n = 17); HbA1c 34.9 ± 2.7 to 33.9 ± 2.3 mmol/mol (p = 0.153) (n = 16); HDL cholesterol 1.6 ± 0.43 mmol/L to 1.5 ± 0.51 mmol/L (p = 0.72) (n = 18); LDL cholesterol 3.0 ± 0.71 mmol/L to 3.6 ± 1.4 mmol/L (p = 0.09) (n = 18); total cholesterol 5.3 ± 0.9 mmol/L to 5.8 ± 1.7 mmol/L (p = 0.21) (n = 18); triglycerides 1.5 ± 0.8 mmol/L to 1.6 ± 0.9 mmol/L (p = 0.98) (n = 16); and CRP 2.4 ± 1.8 mg/L to 2.8 ± 3.6 mg/L (p = 0.738) (n = 15).

Each sample size above reflects the number of participants who had complete data for the respective parameter (variations in sample size across different metabolites are due to missing data, which occurred as a result of incomplete sample collection, processing issues, or participants missing specific tests).

### Serum Metabolomics

Here we report data on glucose, ketones and TCA cycle related metabolites (those that were measurable with our approach) as pre-specified study outcomes.

An exploratory global metabolomics analysis (comprising 358 metabolic features) was performed on a total of 36 serum samples, corresponding to 18 participants who gave a baseline and follow up blood sample, including one baseline non-fasted participant. As expected with human metabolome studies, large variations were noted between the metabolic profiles of individual participants. However, multivariate statistical analysis of the baseline and follow-up sample groups showed a clear separation of the study groups in Partial Least Squares Discriminant Analysis (PLS-DA), suggesting that reliable trends in participant metabolism could be observed within this dataset. For PLS-DA and plots of the metabolomics results please see supplementary figure 2.

In the analysis comparing levels of metabolites between baseline and follow-up, for ketone bodies we observed increases in β-hydroxybutyrate (37563 ± 36445 to 277866 ± 191798 (p = <0.001)), β-ketopentanoate (1548 ± 1367 to 2121 ± 2243 (p = 0.361)) and acetone (5277 ± 7234 to 10776 ± 20957 (p = 0.3)). There were small reductions in acetoacetate (27679 ± 8906 to 20520 ± 12377 (p = 0.054)) and beta-hydroxypentanoate (782 ± 1316 to 34 ± 146 (p = 0.022)). There was a reduction in glucose (210550 ± 188492 to 170700 ± 202785 (p = 0.545)). For TCA cycle related metabolites, we observed a decrease in lactate (171216 ± 171955 to 59291 ± 79476 (p = 0.017)) and glutamine (531128 ± 170513 to 472478 ± 151317 (p = 0.283)), an increase in citrate (940 ± 1927 to 3535 ± 4196 p = 0.023) and isocitrate (2674 ± 7548 to 4079 ± 8837 (p =0.611)) and no change in cis-aconitate (3520 ± 2989 to 3504 ± 3179 to (p = 0.998)).

### Brain Metabolite Levels

The MRS data below includes all usable scans from the 27 participants, including those who did not complete the dietary intervention or attend for follow up (baseline n = 25, follow up n = 19; including one baseline non-fasted scan). MRS data from the RDLPFC was additionally missing for n = 1 participant at both visits since this acquisition was included after the study had begun. n = 6 participants did not attend the follow-up imaging visit. Overall, n = 25 ACC, n = 25 PCC, and n = 24 RDLPFC session one scans and n = 19 ACC, n = 19 PCC, and n = 18 RDLPFC session two scans were acquired.

Automatic quality assessment of the MRS scans resulted in the exclusion of MRS scans of the ACC with linewidth ≥ 0.1ppm from the first (n = 3) and second (n = 1) scanning sessions. No RDLPFC or PCC scans were automatically excluded. Additional spectra were excluded following visual inspection (n = 5 visit 1, n = 6 visit 2), all in the RDLPFC. Creatine and water line widths for included spectra were 6.30±1.80 and 6.55±0.88 Hz, respectively. After automatic and manual exclusions, good fit was achieved in n = 22 ACC, n = 25 PCC, and n = 19 RDLPFC session one scans and n = 18 ACC, n = 19 PCC, and n = 12 RDLPFC session two scans, and were therefore useable in the descriptive analysis. Additionally, n = 16 ACC, n = 19 PCC, and n = 11 RDLPFC paired data sets were useable for calculating paired t-tests.

Neurometabolite concentration estimates are reported in Table 1. Glx (glutamate+glutamine) decreased from baseline in both the ACC (concentration change: -1.50 ± 2.40 [-2.78, -0.22] mM) and PCC (-2.14 ± 1.94 [-3.08, -1.21] mM). A small decrease in total choline was observed in both the ACC ( -0.17 ± 0.20 [-0.28, -0.06] mM) and PCC (-0.12 ± 0.12 [-0.18, -0.06] mM) and a decrease in myoinositol was observed in the PCC (-0.39 ± 0.69 [-0.72, -0.06] mM). No significant differences on any of the metabolites were observed in the RDLPFC.

**Table 1.**
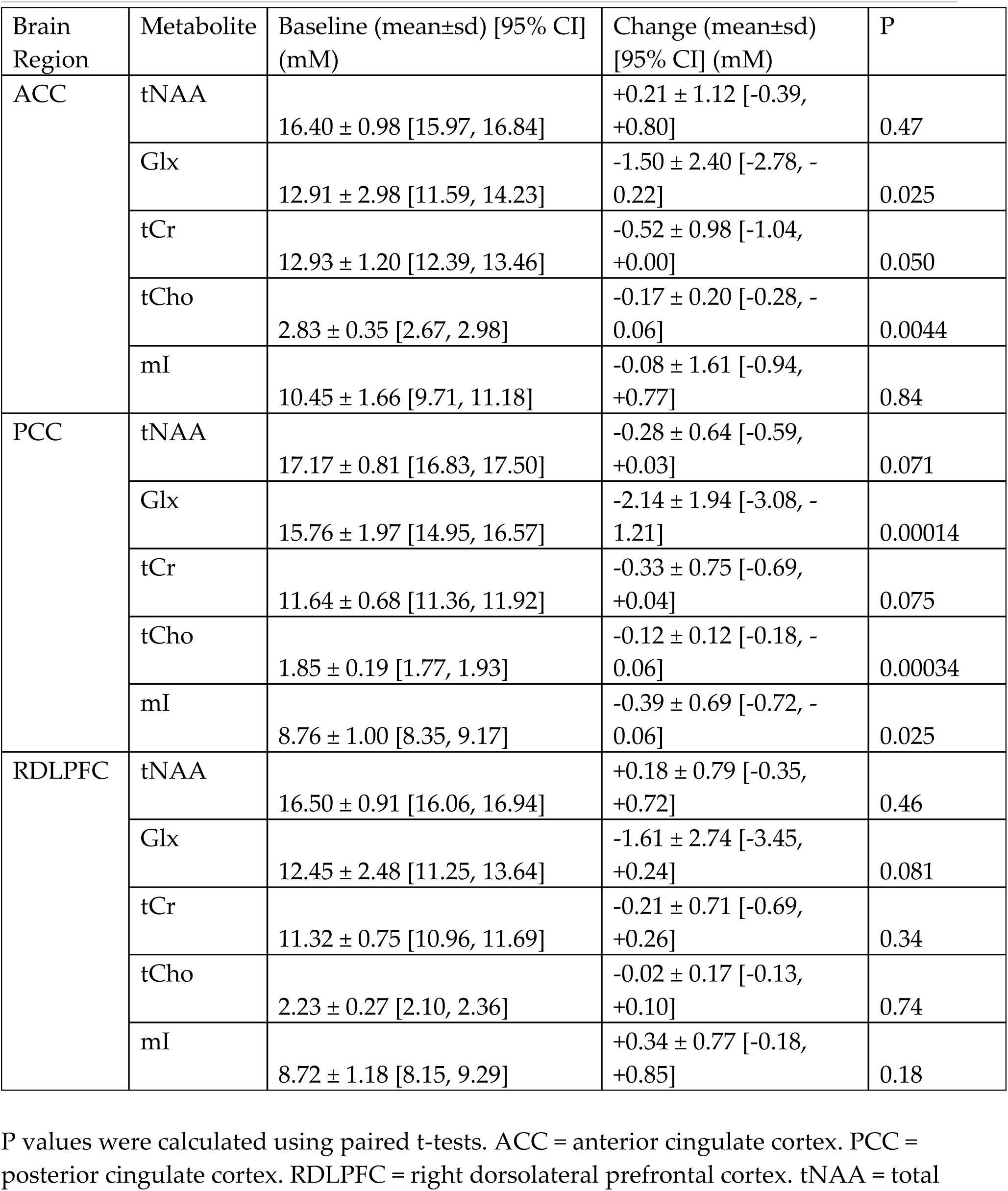
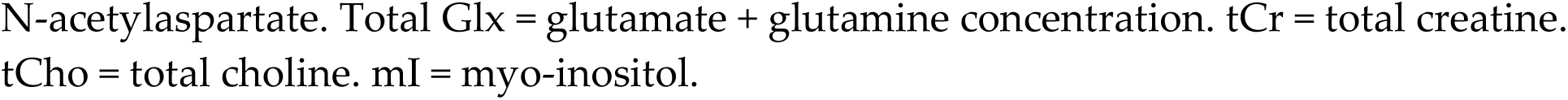
Mean metabolite concentrations estimated using MR spectroscopy before and after ketogenic diet intervention.

### Actigraphy

The actigraphy data had low completion rates with only 65% of the expected data returned for the 20 participants completing the study.(14) The available data derived from actigraphy had substantial day-to-day variability both within and between subjects, precluding any meaningful assessment of the effect of the diet intervention on sleep and/or rest-activity rhythmicity.

## DISCUSSION

Here we report pilot study findings on the short-term effect of a modified ketogenic diet on clinical, metabolic and brain MRS outcomes in euthymic participants with bipolar disorder. Overall, we found preliminary evidence that ketone levels in the blood may be correlated with positive changes in daily mood, energy, impulsivity and anxiety (but not speed of thought). Level of ketosis is considered to be important for seizure reduction in epilepsy with higher levels correlated with seizure control. (21)

There was a positive effect of the ketogenic diet on some cardiometabolic parameters, with improvements in weight and systolic blood pressure, and with no statistically significant effect on lipid profile. These metabolic changes occurred while participants remained on their current medications. One third of the participants completing the study opted out of the dietary cessation period to remain on a ketogenic diet beyond the study period. Similar observations of voluntary, continued adherence post-study are documented in ketogenic diet studies in epilepsy.(22,23)

We also observed preliminary evidence that the ketogenic diet was associated with decreased brain Glx (glutamate plus glutamine concentration) in the ACC and decreased Glx and myoinositol in the PCC. Elevated glutamate is among the most consistent findings in brain MRS studies of bipolar disorder (24–26) and glutamate metabolism is proposed as a mechanism of action of several anti-seizure medications shared between epilepsy and bipolar disorder. Our observation of decreased Glx post-intervention is in line with previous research detecting a reduction in glutamate, glutamine, and Glx in people with bipolar disorder who were good responders to pharmacological interventions (27–31) and to total sleep deprivation and light therapy (32,33). We have previously proposed possible roles of glutamate and myoinositol in cerebral metabolism and insulin signalling in bipolar disorder (9,34). These findings are preliminary from a small-scale study but merit further investigation in studies exploring potential biological mechanisms shared between bipolar, antiseizure medications and ketosis.(34)

In the targeted metabolomics analyses of TCA related metabolites, we observed a reduction in serum lactate. Elevated lactate is also a consistent serum biomarker in bipolar disorder indicated by systematic review.(35) It has been proposed as an indicator of mitochondrial dysfunction in bipolar disorder and other neurological conditions which are accompanied by psychiatric symptoms.(36)

As a pilot study to inform the design of a future randomized controlled trial, there are a number of important limitations. Importantly, this study was not powered to demonstrate statistically significant differences in mental health, metabolic or brain MRS outcomes, so many of the findings reported above are preliminary. The limited sample size also means less precise parameter estimates, increased risk of false positive and false negative findings. We have also not corrected for multiple testing. Further, all participants - many of whom were interested in nutritional approaches - were recruited from a bipolar disorder charity and may not therefore be representative of patients recruited from a clinical setting. The design of this study was single-arm with no control group and was unblinded (for both participants and research staff) and so results may be subject to biases. Dietary interventions, such as the ketogenic diet, are not possible to blind in a trial. We present these data as exploratory and requiring replication and further validation within larger future studies.

## CONCLUSIONS

In this pilot study of a ketogenic diet intervention in euthymic individuals with bipolar disorder, we observed some evidence of potential mental health and metabolic benefits. Given the substantial cardiometabolic risk associated with bipolar disorder and the urgent need for new adjunctive (and non-medication) treatment strategies, replication of these findings and a randomized controlled trial are now warranted. Tentatively, our findings suggest that at least a proportion of patients with bipolar disorder may benefit clinically from an adjunctive metabolic treatment approach such as the ketogenic diet.

## Author Contributions

IC, HC, and DJS were responsible for conceptualising the research question. IC, NN, HG, SS, EM, TM, KB, MT, MM-G, JN, AM, CF, HC and DJS were responsible for designing the study. NN, HG, BM, FC were responsible for data collection. IC, HG, IK, SL, SS, GT, MT, MG, TM, KB, TZ, GG, HC and DJS were responsible for analysis and interpretation of the data. IC, MT, MG, TM, KB and DJS were responsible for drafting the paper. All authors were involved in subsequent revisions, and agree to be accountable for all aspects of the work.

## Data Availability Statement

The data that support the findings of this study are not publicly available as explicit consent was not sought from participants.

## Acknowledgements

We would like to thank Stellate Communications for their assistance with the preparation of figures for this manuscript.

## Transparency Declaration

The lead author IC and manuscript guarantor DS affirm that the manuscript is an honest, accurate, and transparent account of the study being reported; that no important aspects of the study have been omitted; and that any discrepancies from the study as planned have been explained.

