## Supplementary Materials for "A Pilot Study of a Ketogenic Diet in Bipolar Disorder: Clinical, Metabolic and Magnetic Resonance Spectroscopy Findings"

**A Pilot Study of a Ketogenic Diet in Bipolar Disorder: Clinical, Metabolomic and Magnetic Resonance Spectroscopy Outcomes Supplementary Materials**

### Supplementary Metabolomics Methods

Metabolites were extracted from 25 µL serum by adding 1000 µL of ice-cold chloroform/methanol/water (1/3/1 ratio) and mixing vigorously on a vortex for 5 min at 4 °C. The extraction mixtures were centrifuged at 4 °C and 13,000 ×g for 3 min. A 200 µL aliquot of the extracts were transferred into sterile screw-top vials and stored at −80 °C until metabolomics analysis. The untargeted metabolomics analysis was performed using liquid chromatography (LC) coupled to ion mobility (IM) quadrupole time of flight (qTOF) mass spectrometry (MS) as described previously.

The LC-IMS-MS instrumentation used in these experiments consisted of an Agilent 1290 Infinity II series UHPLC system hyphenated with an Agilent 6560 IM-qTOF with a Dual Agilent Jet Stream Electrospray Ionization (ESI) source. In brief, LC-IMS-MS separation was performed on an InfinityLab Poroshell 120 HILIC-Z, 2.1 mm × 50 mm, 2.7 µm UHPLC column (Agilent Technologies 689775-924, Santa Clara, CA) coupled to an InfinityLab Poroshell 120 HILIC-Z, 3.0 mm × 2.7 µm UHPLC guard column (Agilent Technologies 823750-948). A 3.5 min gradient was run using two different solvent systems, composed of low and high pH. Data were acquired in the positive ionisation mode using low pH solvent A (10 mM ammonium formate in water, pH 3) and solvent B (10 mM ammonium formate in water/acetonitrile (1/9), pH 3). Similarly, data were acquired in the negative ionisation mode using high pH solvent A (10 mM ammonium acetate in water, pH 9) and solvent B (10 mM ammonium acetate in water/acetonitrile (1/9), pH 9). The solvent gradient for both positive and negative ionisation modes consisted of 93% solvent B at the start of the run, which was reduced to 80% in 1.8 min, and further to 70% in 0.2 min, where it was maintained for 0.3 min. Subsequently, the column was returned to the initial condition of 93% of solvent B at 2.35 min and maintained until 3.5 min. The column was kept at a constant temperature of 30 °C and a constant flow rate of 0.8 mL/min was maintained during the entire chromatographic separation. For analysis, 1 µL of sample was injected into the column. A pooled quality control sample was generated by combining equal volumes of each sample and this was injected five times at the beginning of the experiment to condition the column and after every five samples to monitor the instrument state over the course of data acquisition. Data were acquired in the 50-1700 m/z range, with an MS acquisition rate of 0.8 frames/s for both ionisation modes. The nebulizer pressure was set to 60 psi, gas temperature to 225 °C and drying gas (N2) flow to 13 L/min. Sheath gas was set to 340 °C with a flow rate of 12 L/min, and the instrument was operated at a capillary voltage of 3000 V, nozzle voltage of 200 V, fragmentor voltage of 395 V, and octupole voltage of 750 V. Instrument calibration and tuning was performed separately for each ionisation polarity using the ESI-L low concentration tuning mix from Agilent Technologies. A reference mass solution consisting of 50 µM ammonium trifluoroacetate, 5 µM purine and 1.125 µM HP-0921 was injected continuously into each sample to recalibrate for accurate mass and drift time during data processing. The ES-TOF reference mass solution kit was purchased from Agilent Technologies (Santa Clara, CA).

The Agilent MassHunter 10.0 software suite was used for data acquisition and data processing. Briefly, ion-mobility multiplexed data files and calibration files obtained from the MassHunter Data Acquisition 10.0 software were demultiplexed using the PNNL PreProcessor v2020.03.23. The default settings were applied to the data for demultiplexing, moving average smoothing, saturation repair, and spike removal. Data files were then recalibrated for accurate mass and 6 drift time using the AgtTofReprocessUi and IM-MS Browser 10.0, respectively. Two reference masses, m/z 121.050873 and 922.009798 (positive ionisation mode) and m/z 112.985587 and 1033.988109 (negative ionisation mode) were used for recalibration. Mass Profiler 10.0 was used to extract molecular features with a retention time tolerance of 0.3 min, drift time tolerance of 1.5% and accurate mass tolerance of (5 ppm + 2 mDa). Additional peak deconvolution was performed in the High Resolution Demultiplexer (HRdm) 1.0 beta v41 using the raw multiplexed data, the reconstructed demultiplexed data, and the Mass Profiler features lists (.cef files). Molecular features were re-extracted from HRdm files using Mass Profiler 10.0 and annotated using accurate mass and CCS values with the McLean CCS Compendium PCDL (version 20191101). A 10 ppm window was applied for m/z matches, a CCS value tolerance of 1% was applied, and the positive ion species (M+H)+, (M+Na)+ and the negative ion species (M-H)- were searched. Metabolite annotations are therefore classified as level 2 according to the metabolite standards initiative.

Multivariate statistical analysis and pathway enrichment analysis were performed using the MetaboAnalyst 5.0 web-based platform. The data were log-transformed and pareto-scaled to generate partial least squares - discriminant analysis (PLS-DA) plots and heatmaps. For a more detailed analysis of the altered pathways, the annotated features with their relative intensities was submitted to the pathway analysis tool, log-transformed, pareto scaled and examined against the Homo sapiens KEGG pathway library, using global test and relative betweenness centrality methods. Peak intensities of paired samples from individual participants were also plotted using excel to better understand trends within the study groups.

**Supplementary Figure 1 Metabolomics Principal Component Analysis**

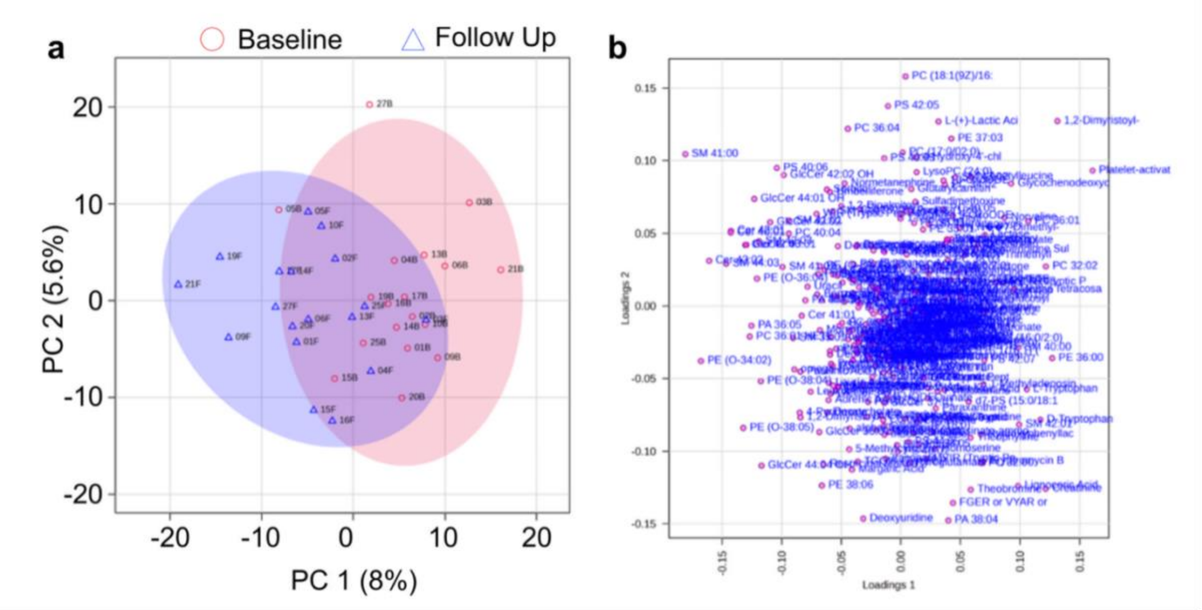


(a) Principal Component Analysis (PCA) score plot shows the serum metabolome at baseline (red circle) and follow up (blue triangle). The first and second components explain 8 and 5.6% of the variation, respectively. (b) The loadings plot corresponding to the PCA shows the original variables (N = 18).

### Supplementary Figure 2 Metabolomics Results

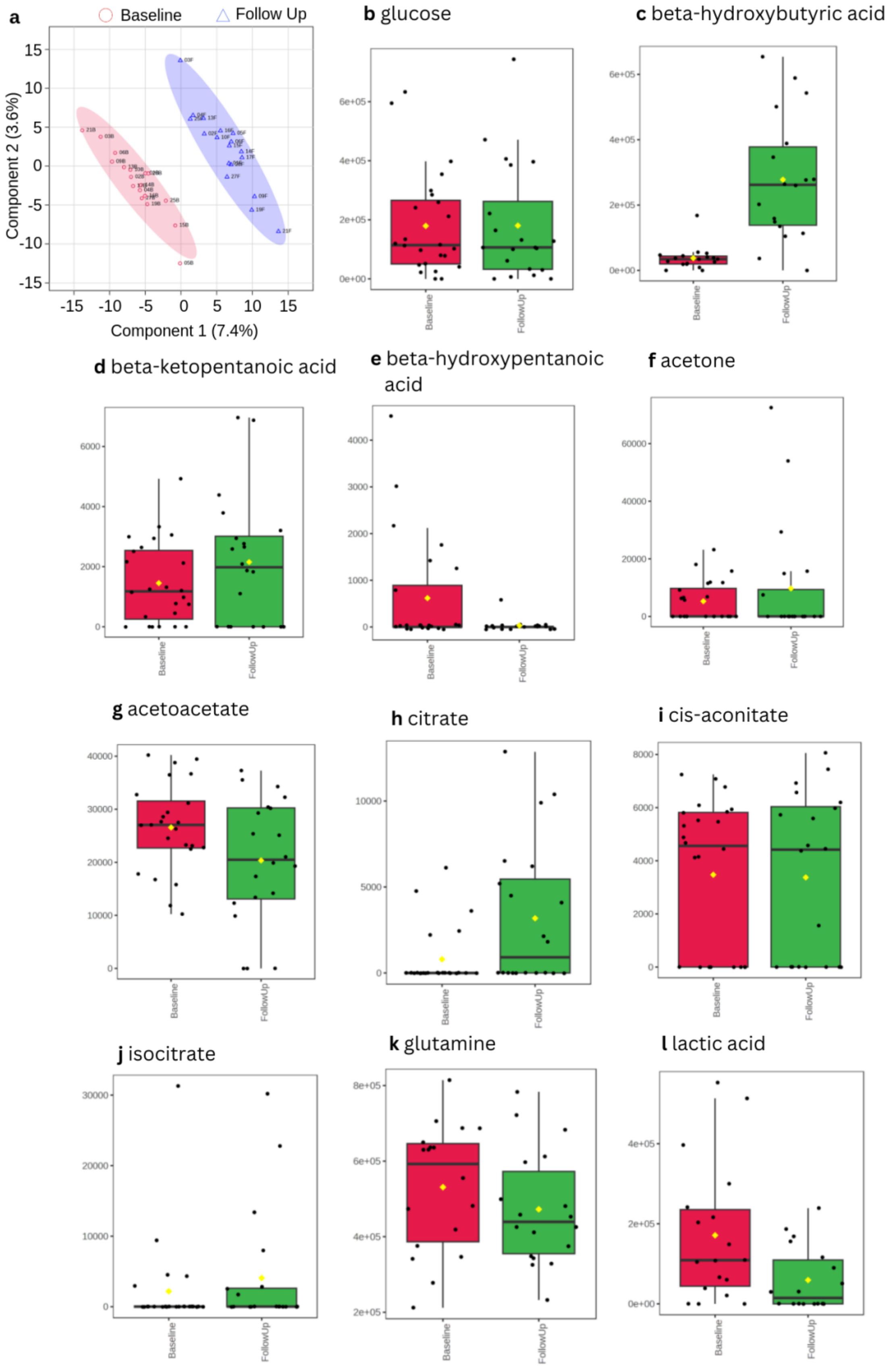

(a) Partial Least Squares Discriminant Analysis (PLS-DA) score plot shows separation of the serum metabolome at baseline (red circle) and follow up (blue triangle). The first and second components explain 7.4 and 3.6% of the variation, respectively. (b) glucose (c -g) ketone bodies (h -l) TCA cycle related metabolites associated with ketogenic diet.

**MRS Quality Assessment Supplementary Methods**

An automatic quality assessment of the MRS scans resulted in the exclusion of MRS scans of the ACC with high linewidth from the first (N=3) and second (N=1) scanning sessions. No RDLPFC or PCC scans were automatically excluded. Additional spectra were excluded following visual inspection (N=5 visit 1, N=6 visit 2), all in the RDLPFC. Creatine and water line widths for included spectra were 6.30±1.80 and 6.55±0.88 Hz, respectively. Figure 4 shows representative acquired and fitted model spectra from a single participant. After automatic and manual exclusions, good fit was achieved in N=22 ACC, N=25 PCC, and N=19 RDLPFC session one scans and N=18 ACC, N=19 PCC, and N=12 RDLPFC session two scans, and were therefore useable in the descriptive analysis. Additionally, N=16 ACC, N=19 PCC, and N=11 RDLPFC paired data sets were useable for calculating paired t-tests.

**MRS Supplementary Table 1 Data for all Fitted Signals**

| Brain Region | Metabolite | Baseline (mean±sd) [95% CI] (mM) | Change (mean±sd) [95% CI] (mM) |
|---|---|---|---|
| ACC | Asc | 0.86 ± 0.69 [0.56, 1.17] | -0.27 ± 1.06 [-0.84, +0.29] |
| | Asp | 2.31 ± 1.08 [1.84, 2.79] | -0.38 ± 0.97 [-0.90, +0.13] |
| | Cr | 5.69 ± 1.45 [5.05, 6.33] | +0.02 ± 1.39 [-0.73, +0.76] |
| | CrCH2 | 1.30 ± 0.94 [0.88, 1.72] | +0.49 ± 0.86 [+0.03, +0.95] |
| | GABA | 4.08 ± 1.59 [3.38, 4.79] | +1.39 ± 2.64 [-0.02, +2.79] |
| | GPC | 1.70 ± 0.58 [1.45, 1.96] | -0.39 ± 0.43 [-0.62, -0.16] |
| | GSH | 2.36 ± 0.36 [2.20, 2.52] | -0.31 ± 0.63 [-0.65, +0.02] |
| | Gln | 1.05 ± 1.26 [0.50, 1.61] | -0.58 ± 1.14 [-1.18, +0.03] |
| | Glu | 11.91 ± 2.00 [11.02, 12.79] | -0.93 ± 1.63 [-1.79, -0.06] |
| | mI | 10.45 ± 1.66 [9.71, 11.18] | -0.08 ± 1.61 [-0.94, +0.77] |
| | Lac | 1.58 ± 0.76 [1.25, 1.92] | -0.33 ± 0.65 [-0.68, +0.02] |
| | NAA | 14.80 ± 0.91 [14.40, 15.20] | +0.40 ± 0.96 [-0.11, +0.91] |
| | NAAG | 1.52 ± 0.40 [1.34, 1.70] | -0.21 ± 0.55 [-0.50, +0.09] |
| | PCh | 1.17 ± 0.50 [0.95, 1.40] | +0.22 ± 0.48 [-0.04, +0.48] |
| | PCr | 7.23 ± 1.82 [6.43, 8.04] | -0.54 ± 1.82 [-1.51, +0.44] |
| | PE | 2.73 ± 1.16 [2.22, 3.25] | -0.41 ± 1.26 [-1.07, +0.26] |
| | sI | 0.61 ± 0.30 [0.48, 0.75] | -0.10 ± 0.19 [-0.21, -0.00] |

| | | | |
|---|---|---|---|
| | **Tau** | 5.59 ± 1.12 [5.09, 6.09] | +0.24 ± 1.48 [-0.55, +1.02] |
| | **MM09** | 2.07 ± 0.56 [1.82, 2.32] | -0.02 ± 0.59 [-0.33, +0.30] |
| | **MM12** | 2.81 ± 0.79 [2.46, 3.16] | +0.26 ± 1.03 [-0.29, +0.81] |
| | **MM14** | 1.90 ± 1.15 [1.39, 2.41] | +0.12 ± 1.24 [-0.54, +0.78] |
| | **MM17** | 2.60 ± 1.80 [1.80, 3.39] | -0.89 ± 1.85 [-1.87, +0.10] |
| | **MM20** | 2.44 ± 0.94 [2.02, 2.85] | -0.51 ± 1.10 [-1.10, +0.07] |
| | **Lip09** | 5.12 ± 0.77 [4.78, 5.45] | +0.20 ± 0.90 [-0.28, +0.68] |
| | **Lip13** | 1.77 ± 1.05 [1.30, 2.23] | -0.13 ± 1.41 [-0.88, +0.62] |
| | **Lip20** | 2.47 ± 0.56 [2.22, 2.72] | -0.32 ± 0.73 [-0.71, +0.07] |
| | **tNAA** | 16.40 ± 0.98 [15.97, 16.84] | +0.21 ± 1.12 [-0.39, +0.80] |
| | **Glx** | 12.91 ± 2.98 [11.59, 14.23] | -1.50 ± 2.40 [-2.78, -0.22] |
| | **tCho** | 2.83 ± 0.35 [2.67, 2.98] | -0.17 ± 0.20 [-0.28, -0.06] |
| | **tCr** | 12.93 ± 1.20 [12.39, 13.46] | -0.52 ± 0.98 [-1.04, +0.00] |
| **PCC** | **Asc** | 0.17 ± 0.35 [0.02, 0.31] | +0.06 ± 0.37 [-0.12, +0.24] |
| | **Asp** | 2.97 ± 0.80 [2.64, 3.29] | -0.14 ± 0.78 [-0.52, +0.24] |
| | **Cr** | 7.10 ± 0.65 [6.83, 7.37] | -0.34 ± 1.13 [-0.89, +0.20] |
| | **CrCH2** | 1.51 ± 0.48 [1.31, 1.71] | +0.02 ± 0.36 [-0.15, +0.20] |
| | **GABA** | 2.18 ± 0.93 [1.79, 2.56] | +0.20 ± 1.04 [-0.30, +0.70] |
| | **GPC** | 1.14 ± 0.25 [1.04, 1.24] | -0.22 ± 0.35 [-0.39, -0.06] |
| | **GSH** | 1.95 ± 0.30 [1.82, 2.07] | -0.17 ± 0.26 [-0.30, -0.05] |
| | **Gln** | 2.93 ± 1.12 [2.46, 3.39] | -1.11 ± 1.13 [-1.65, -0.57] |
| | **Glu** | 12.88 ± 1.09 [12.43, 13.34] | -1.04 ± 1.01 [-1.52, -0.55] |
| | **mI** | 8.76 ± 1.00 [8.35, 9.17] | -0.39 ± 0.69 [-0.72, -0.06] |
| | **Lac** | 1.34 ± 1.32 [0.79, 1.88] | +0.21 ± 2.76 [-1.12, +1.54] |
| | **NAA** | 15.13 ± 0.76 [14.82, 15.45] | -0.07 ± 0.77 [-0.44, +0.30] |
| | **NAAG** | 1.96 ± 0.42 [1.79, 2.14] | -0.22 ± 0.43 [-0.42, -0.01] |
| | **PCh** | 0.74 ± 0.23 [0.65, 0.84] | +0.10 ± 0.30 [-0.04, +0.25] |
| | **PCr** | 4.54 ± 0.82 [4.20, 4.87] | +0.02 ± 1.34 [-0.63, +0.66] |
| | **PE** | 2.26 ± 0.68 [1.98, 2.54] | +0.28 ± 0.94 [-0.17, +0.74] |
| | **sI** | 0.48 ± 0.23 [0.38, 0.58] | +0.02 ± 0.14 [-0.05, +0.09] |
| | **Tau** | 4.63 ± 0.79 [4.31, 4.96] | +0.20 ± 0.95 [-0.25, +0.66] |
| | **MM09** | 3.27 ± 0.88 [2.90, 3.63] | -0.12 ± 1.17 [-0.68, +0.44] |
| | **MM12** | 2.14 ± 1.18 [1.66, 2.63] | +0.71 ± 1.44 [+0.01, +1.40] |
| | **MM14** | 2.25 ± 1.52 [1.63, 2.88] | +0.64 ± 2.79 [-0.70, +1.99] |
| | **MM17** | 4.02 ± 2.71 [2.90, 5.14] | -0.84 ± 2.89 [-2.23, +0.56] |
| | **MM20** | 4.28 ± 0.95 [3.89, 4.67] | -0.53 ± 1.16 [-1.09, +0.03] |
| | **Lip09** | 5.90 ± 1.17 [5.42, 6.38] | -0.06 ± 1.21 [-0.64, +0.52] |
| | **Lip13** | 0.88 ± 1.08 [0.43, 1.32] | +1.08 ± 2.08 [+0.08, +2.08] |
| | **Lip20** | 2.43 ± 0.70 [2.15, 2.72] | -0.51 ± 0.60 [-0.80, -0.22] |

| | | | |
|---|---|---|---|
| | **tNAA** | 17.17 ± 0.81 [16.83, 17.50] | -0.28 ± 0.64 [-0.59, +0.03] |
| | **Glx** | 15.76 ± 1.97 [14.95, 16.57] | -2.14 ± 1.94 [-3.08, -1.21] |
| | **tCho** | 1.85 ± 0.19 [1.77, 1.93] | -0.12 ± 0.12 [-0.18, -0.06] |
| | **tCr** | 11.64 ± 0.68 [11.36, 11.92] | -0.33 ± 0.75 [-0.69, +0.04] |
| **RDLPFC** | **Asc** | 0.38 ± 0.41 [0.19, 0.58] | +0.12 ± 0.54 [-0.24, +0.48] |
| | **Asp** | 2.51 ± 0.52 [2.25, 2.76] | -0.30 ± 0.43 [-0.59, -0.01] |
| | **Cr** | 6.95 ± 1.31 [6.32, 7.58] | -0.31 ± 1.21 [-1.12, +0.50] |
| | **CrCH2** | 1.19 ± 0.73 [0.83, 1.54] | -0.28 ± 0.70 [-0.75, +0.19] |
| | **GABA** | 2.11 ± 1.04 [1.61, 2.61] | +0.50 ± 1.16 [-0.28, +1.28] |
| | **GPC** | 1.52 ± 0.43 [1.32, 1.73] | +0.05 ± 0.36 [-0.20, +0.29] |
| | **GSH** | 2.03 ± 0.33 [1.87, 2.19] | -0.08 ± 0.29 [-0.28, +0.11] |
| | **Gln** | 1.94 ± 1.61 [1.16, 2.71] | -1.10 ± 1.80 [-2.31, +0.11] |
| | **Glu** | 10.55 ± 1.10 [10.02, 11.08] | -0.51 ± 1.34 [-1.41, +0.39] |
| | **mI** | 8.72 ± 1.18 [8.15, 9.29] | +0.34 ± 0.77 [-0.18, +0.85] |
| | **Lac** | 1.39 ± 1.00 [0.90, 1.87] | +0.15 ± 0.81 [-0.39, +0.69] |
| | **NAA** | 14.62 ± 0.97 [14.15, 15.08] | +0.25 ± 0.75 [-0.25, +0.76] |
| | **NAAG** | 1.81 ± 0.44 [1.60, 2.03] | -0.08 ± 0.52 [-0.42, +0.27] |
| | **PCh** | 0.74 ± 0.41 [0.55, 0.94] | -0.06 ± 0.33 [-0.28, +0.16] |
| | **PCr** | 4.37 ± 1.34 [3.72, 5.02] | +0.09 ± 1.50 [-0.91, +1.10] |
| | **PE** | 2.57 ± 0.75 [2.21, 2.93] | -0.24 ± 1.09 [-0.98, +0.49] |
| | **sI** | 0.51 ± 0.20 [0.42, 0.61] | -0.06 ± 0.13 [-0.15, +0.03] |
| | **Tau** | 4.47 ± 0.70 [4.13, 4.80] | +0.41 ± 0.71 [-0.07, +0.89] |
| | **MM09** | 2.90 ± 0.65 [2.58, 3.21] | -0.42 ± 0.74 [-0.92, +0.07] |
| | **MM12** | 1.73 ± 1.07 [1.21, 2.24] | -0.56 ± 1.60 [-1.64, +0.52] |
| | **MM14** | 2.26 ± 1.62 [1.48, 3.05] | +1.01 ± 1.87 [-0.24, +2.27] |
| | **MM17** | 3.90 ± 2.87 [2.52, 5.28] | -1.88 ± 3.13 [-3.98, +0.23] |
| | **MM20** | 3.40 ± 0.93 [2.95, 3.84] | -0.43 ± 0.92 [-1.05, +0.19] |
| | **Lip09** | 4.52 ± 1.14 [3.97, 5.07] | +0.45 ± 1.73 [-0.71, +1.61] |
| | **Lip13** | 1.28 ± 1.54 [0.54, 2.03] | -0.25 ± 2.13 [-1.68, +1.18] |
| | **Lip20** | 1.80 ± 0.61 [1.50, 2.09] | -0.20 ± 0.96 [-0.85, +0.44] |
| | **tNAA** | 16.50 ± 0.91 [16.06, 16.94] | +0.18 ± 0.79 [-0.35, +0.72] |
| | **Glx** | 12.45 ± 2.48 [11.25, 13.64] | -1.61 ± 2.74 [-3.45, +0.24] |
| | **tCho** | 2.23 ± 0.27 [2.10, 2.36] | -0.02 ± 0.17 [-0.13, +0.10] |
| | **tCr** | 11.32 ± 0.75 [10.96, 11.69] | -0.21 ± 0.71 [-0.69, +0.26] |

P values were calculated using paired t-tests. ACC = anterior cingulate cortex. PCC = posterior cingulate cortex. RDLPFC = right dorsolateral prefrontal cortex. NAA = total N-acetylaspartate. Glx = glutamate + glutamine concentration. tCr = total creatine. tCho = total choline. mI = myo-inositol. Asc = Ascorbate. Asp = Aspartate. Cr = Creatine. CrCH2 = Creatine methylene. GABA = Gamma-aminobutyric acid. GPC = Glycerophosphocholine. GSH = Glutathione. Gln = Glutamine. Glu = Glutamate. mI = Myo-inositol. Lac = Lactate. NAA = N-acetylaspartate. NAAG = N-acetylaspartylglutamate. PCh = Phosphocholine. PCr = Phosphocreatine. PE = Phosphoethanolamine. sI = Scyllo-Inositol. Tau = Taurine. MM09 = Macromolecule at 0.9ppm. MM12 = Macromolecule at 1.2ppm. MM14 = Macromolecule at 1.4ppm. MM17 = Macromolecule at 1.7ppm. MM20 = Macromolecule at 2.0ppm. Lip09 = Lipid at 0.9ppm. Lip13 = Lipid at 1.3ppm. Lip20 = Lipid at 2.0ppm. tNAA = Total N-acetylaspartate (includes both NAA and NAAG). tCho = Total choline (includes phosphocholine and glycerophosphocholine). tCr = Total creatine (includes creatine and phosphocreatine).

**Supplementary Table 2 MRS Results Expressed as Signal Ratios to the Total Creatine Signal**

| Brain Region | Metabolite | Baseline (mean±sd) [95% CI] (mM) | Change (mean±sd) [95% CI] (mM) | P |
|---|---|---|---|---|
| **ACC** | **tNAA** | 1.361 ± 0.117 [1.309, 1.413] | +0.070 ± 0.161 [-0.015, +0.156] | 0.10 |
| | **Glx** | 0.995 ± 0.187 [0.912, 1.078] | -0.079 ± 0.153 [-0.160, +0.002] | 0.056 |
| | **tCho** | 0.250 ± 0.022 [0.240, 0.259] | -0.005 ± 0.015 [-0.013, +0.003] | 0.18 |
| | **mI** | 0.907 ± 0.117 [0.855, 0.960] | +0.031 ± 0.106 [-0.025, +0.087] | 0.26 |
| **PCC** | **tNAA** | 1.577 ± 0.100 [1.536, 1.618] | +0.017 ± 0.091 [-0.027, +0.061] | 0.43 |
| | **Glx** | 1.358 ± 0.182 [1.284, 1.433] | -0.153 ± 0.177 [-0.238, -0.068] | 0.0014 |
| | **tCho** | 0.182 ± 0.015 [0.175, 0.188] | -0.007 ± 0.010 [-0.011, -0.002] | 0.0095 |
| | **mI** | 0.845 ± 0.077 [0.813, 0.876] | -0.013 ± 0.043 [-0.034, +0.008] | 0.21 |
| **RDLPFC** | **tNAA** | 1.561 ± 0.151 [1.488, 1.633] | +0.043 ± 0.114 [-0.034, +0.120] | 0.24 |
| | **Glx** | 1.103 ± 0.211 [1.002, 1.205] | -0.125 ± 0.208 [-0.265, +0.015] | 0.075 |
| | **tCho** | 0.225 ± 0.025 [0.213, 0.237] | +0.002 ± 0.017 [-0.010, +0.014] | 0.70 |
| | **mI** | 0.865 ± 0.111 [0.812, 0.919] | +0.048 ± 0.086 [-0.010, +0.106] | 0.095 |

*P* values were calculated using paired *t*-tests. ACC = anterior cingulate cortex. PCC = posterior cingulate cortex. RDLPFC = right dorsolateral prefrontal cortex. NAA = total N-acetylaspartate. Total Glx = glutamate + glutamine concentration. tCr = total creatine. tCho = total choline. mI = myo-inositol.

**EMA Instructions v1 (Participants 1-12)**

Dear participant,

Please could you send us the following results from this afternoon as close to 4pm as possible:

Ketone level
Time ketone level taken
Glucose level
Time glucose level taken

Please rate the following out of 1 to 100 at the time of sending your reply:

A) Mood (1 is sad and 100 is happy)
B) Energy levels (1 is low, 100 is high)
C) Speed of thought (1 is slow, 100 is fast)
D) Impulsivity (1 is low, 100 is high)
E) Anxiety (1 is low, 100 is high)

**EMA Instructions v2 (Participants 13-27)**

Dear participant,

Please could you send us the following results from this afternoon as close to 4pm as possible:

Ketone level
Time ketone level taken
Glucose level
Time glucose level taken

Please rate the following from 1 to 100 at the time of sending your reply:

A) Mood
B) Energy levels

C) Speed of thought
D) Impulsivity
E) Anxiety

Please rate the following out of 1 to 100 at the time of sending your reply:

A) Mood (1 indicates your mood being the lowest it has ever been and 100 the highest it has ever been. If your mood feels within a healthy range for you, please rate it between 30-70. If you feel your mood is depressed, please rate it between 0-30. If you feel you are hypomanic or manic, please rate your mood between 70-100)

B) Energy levels (1 indicates your energy being the lowest it has ever been and 100 the highest it has ever been. If your energy levels feel within a healthy range for you, please rate them between 30-70. If you feel your energy levels are lower than they should be, please rate them between 0-30. If you feel they are higher than they should be, please rate them between 70-100)

C) Speed of thought (1 indicates your thoughts are the slowest they have ever been, 100 the fastest they have ever been. If your thought speed feels within a healthy range for you, please rate it between 30-70. If your thought speed feels much slower than it should be, please rate it between 0-30. If you feel it is much faster than it should be, please rate it between 70-100. 10

D) Impulsivity (1 indicates the least impulsive you have ever felt, 100 indicates the most impulsive you have ever felt. If you do not feel impulsive, please score between 0-25, for mild impulsivity please score between 25-50, for moderate impulsivity please score between 50-75, and for extreme impulsivity please score between 75-100).

E) Anxiety (1 indicates the least anxious you have ever felt, 100 indicates the most anxious you have ever felt. If you do not feel anxious, please score between 0-25, for mild anxiety please score between 25-50, for moderate anxiety please score between 50-75, and for extreme anxiety please score between 75-100).
